# A practical strategy for data assimilation of cerebral intra-aneurysmal flows using a variational method with boundary control of velocity

**DOI:** 10.1101/2024.05.24.24307838

**Authors:** Tsubasa Ichimura, Shigeki Yamada, Yoshiyuki Watanabe, Hiroto Kawano, Satoshi Ii

## Abstract

**Background and objective:** Evaluation of hemodynamics is crucial to predict growth and rupture of cerebral aneurysms. Variational data assimilation (DA) is a powerful tool to characterize patient-specific intra-aneurysmal flows. The DA inversely estimates a boundary condition in fluid equations using personalized flow data; however, its high computational cost in optimization problems makes its use impractical.

**Methods:** This study proposes a practical strategy for the DA to evaluate patient-specific intra-aneurysmal flows. To estimate personalized flows, a variational DA was combined with computational fluid dynamics (CFD) and four-dimensional flow magnetic resonance imaging (4D flow MRI) for intra-aneurysmal velocity data, and an inverse problem was solved to estimate the spatiotemporal velocity profile at a boundary of the aneurysm neck. To circumvent an ill-posed inverse problem, model order reduction based on a Fourier series expansion was used to describe temporal changes in state variables.

**Results:** In numerical validation using synthetic data from the CFD, the present DA achieved excellent agreement with the CFD as ground truth, with velocity mismatch within the 4%-7% range. In flow estimations for three patient-specific datasets, the proposed DA shows the velocity mismatch within the 35%-63% range, which is less than half that of the CFD using main vessel branches, and would mitigate unphysical velocity distributions in the 4D flow MRI.

**Conclusions:** By focusing only on the intra-aneurysmal region, the present strategy based on the DA provides an attractive way to evaluate personalized flows in aneurysms with greater reliability than conventional CFD and better efficiency than existing DA approaches.

## 1. Introduction

A cerebral aneurysm is a cerebrovascular disorder in which part of vessel wall develops an outward bulge. Rupture of a cerebral aneurysm is a major factor in subarachnoid hemorrhage and has a mortality rate of up to 35% [1], although the rupture rate is only approximately 2% per year [2]. Since there is a trade-off between the risks of rupture and clinical intervention, a high-level evaluation to predict the rupture risk is highly desirable.

It has been argued that growth and rupture of an aneurysm are generated by a three-way relationship among the pathobiology of the wall, aneurysmal geometry, and intra-aneurysmal flow [3,4]. To quantify the hemodynamics of cerebral aneurysms, phase-contrast magnetic resonance imaging (MRI), also termed four-dimensional (4D) flow MRI, and computational fluid dynamics (CFD) have been conducted and compared [5-9]. However, it remains challenging to adequately quantify hemodynamic features in vivo because the spatiotemporal resolution of 4D flow MRI is limited and it is difficult to set patient-specific boundary conditions for CFD [10].

Data assimilation (DA) is a powerful tool to quantify time-dependent intra-aneurysmal flows in each patient. It predicts a more statistically reliable velocity field based on the observed and modelled velocities from 4D flow MRI and CFD, respectively. In particular, an inverse approach is preferably used to strictly satisfy physical constraints [11]. Several types of inverse DA algorithms have been developed for steady and time-dependent problems; these algorithms estimate boundary conditions (e.g., the inlet/outlet velocity or surface traction) or flow velocity field, using a feedback control method [12], variational method [13-17], and sequential (Kalman filter-based) method [18]. However, these DA approaches analyze the main vessels in addition to an aneurysm, resulting in a high computational cost with respect to the computation time and memory requirement for such time-dependent problems. Moreover, since solutions are often sensitive to material parameters such as blood viscosity and vessel shape reconstructed from image-segmentation, uncertainty estimates under many different analysis conditions are required for each patient, which further increases the computational cost. These difficulties should be mitigated so that DA can be used to quantify patient-specific intra-aneurysmal flows.

This study aimed to develop a practical strategy for the DA to evaluate patient-specific intra-aneurysmal flows. Using a variational DA coupled with CFD and measured velocity data, and limiting the domain of analysis to the inside of the aneurysm, an inverse problem was solved to estimate the spatiotemporal velocity profile at the aneurysm neck. Furthermore, to circumvent an ill-posed inverse problem, model order reduction based on a Fourier series expansion was employed to describe temporal changes in state variables. The model was validated numerically using synthetic data obtained from CFD with given boundary conditions, and then the model feasibility was investigated by applying it to three patient-specific MRI datasets.

## 2. Material and methods

### 2.1. Patient-specific data

Time-of-flight magnetic resonance angiography (TOF-MRA) and 4D flow MRI were conducted using three patients, each of whom had an aneurysm at an internal carotid artery. The study was approved by the ethics committees for human research at Shiga University of Medical Science (IRB Number: R2019-227). The TOF-MRA images (pixel size, 0.3906 mm; slice thickness, 0.6 mm) were used for reconstruction of vessel geometries. The spatiotemporal velocity fields of the blood stream were reconstructed from 4D flow MRI data (pixel size, 0.7031 mm; slice thickness, 1.0 mm; temporal resolution, 12 frames per heartbeat) acquired using velocity encoding (VENC) at 40 cm/s (VENC40) and 120 cm/s (VENC120).

Extraction and smoothing of vessel shapes were performed using Mimics (Materialise, Belgium) and Meshmixer (Autodesk, USA). For further usage, surface shapes of the vessels were implicitly described using a discrete level-set function with a signed distance. The 4D flow MRI and TOF-MRA datasets were co-registered using a rigid transformation based on global geometry information stored in the headers of their corresponding DICOM files. The velocity field was extracted from the 4D flow MRI data using VENC, and the velocity inside the vessel was then extracted using the level-set function for the vessel reconstructed from the TOF-MRA image (Fig. 1). Only velocity vectors at positions greater than one pixel from the vessel wall were extracted to eliminate artifacts located near the wall. See the details of the vessel shapes and aneurysm sizes for each of the three patients (sample A, B and C) in Appendix A.

**FIGURE 1.**
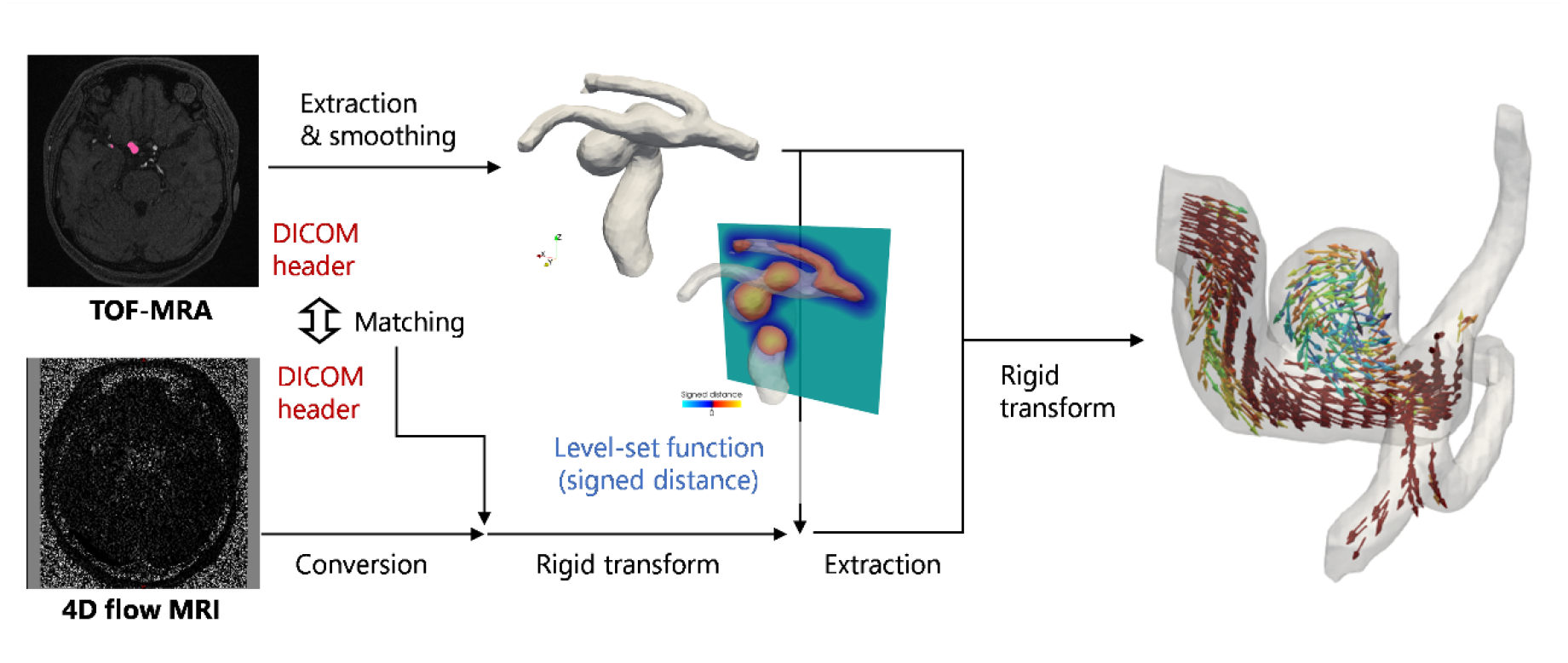
Reconstruction of vessel shape and blood velocity obtained from time-of-flight magnetic resonance angiography (TOF-MRA) and four-dimensional flow magnetic resonance imaging (4D flow MRI).

### 2.2. Application of DA to cerebral aneurysms

#### 2.2.1. 4D variational (4D-Var) method

In this study, a 4D-Var method was applied for DA. In this method, the interior of an aneurysm without branching vessels is introduced as an analysis domain, and an optimization problem is solved to minimize velocity mismatch between the mathematical model and experimental observation in that domain. An optimal control concept is used to solve the problem by imposing a spatiotemporal velocity profile on the aneurysm neck as a boundary condition for the incompressible Navier–Stokes equations, and this velocity profile is inversely estimated as a design variable. The mathematical description can be given as

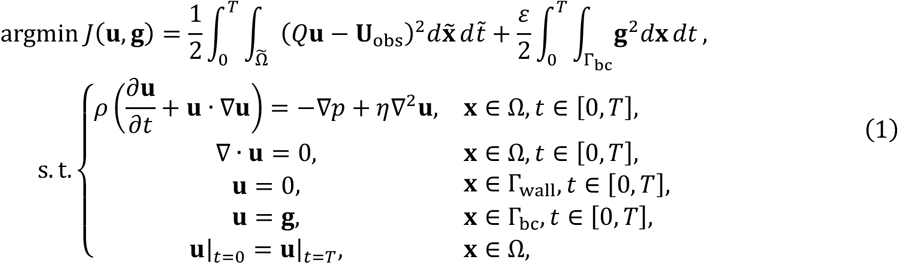

where *J*(**u, g**) is the objective function consisting of two cost functions; the first represents the total data mismatch between the model velocity **u**(**x**, *t*) and observed velocity 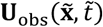 in the spatiotemporal domain comprising the analysis space **x** ∈ Ω, observation space 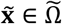 and time *t*, 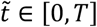, where *T* is the period of pulsatile flow; and the second represents a zeroth-order Tikhonov regularization function with a parameter *ε* for the design variable **g**(**x**, *t*) (i.e., the boundary velocity on the aneurysm neck Γ_bc_) to mitigate sensitivity to data noise and an ill-posed inverse problem. Parameter *Q* is the operator mapping the solution field of the mathematical model to the observation. In Eq. (1), the incompressible Navier– Stokes equations act as constraints for the model velocity **u** by imposing a no-slip condition at the fixed wall Γ_wall_ and an inlet/outlet velocity **g** at the aneurysm neck Γ_bc_. Here, *p*(**x**, *t*) is the fluid pressure, *ρ* is although the fluid density, *η* is the fluid viscosity, and **n** is the unit normal vector on the surface Γ_bc_. In general, although initial conditions are simultaneously estimated in the 4D-Var, the blood flows can be assumed to behave as pulsatile flows with a heart period *T*, and thus, temporal periodicity is imposed in the final condition in Eq. (1), i.e., **u**|_*t*=0_ = **u**|_*t*=*T*_ where **u**|_*t*=0_ = **u**(**x**, 0) and **u**|_*t*=*T*_ = **u**(**x**, *T*), instead of estimating the initial conditions. To satisfy the mass conservation, the flowrate condition 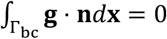 should be satisfied for incompressible flows.

To simplify the numerical setup, the extracted aneurysms were rotated so that the aneurysm extrusion pointed along the *z* direction and the inlet/outlet surface Γ_bc_ was defined to be a cross-section through the neck in the *xy* plane (Fig. 2).

**FIGURE 2.**
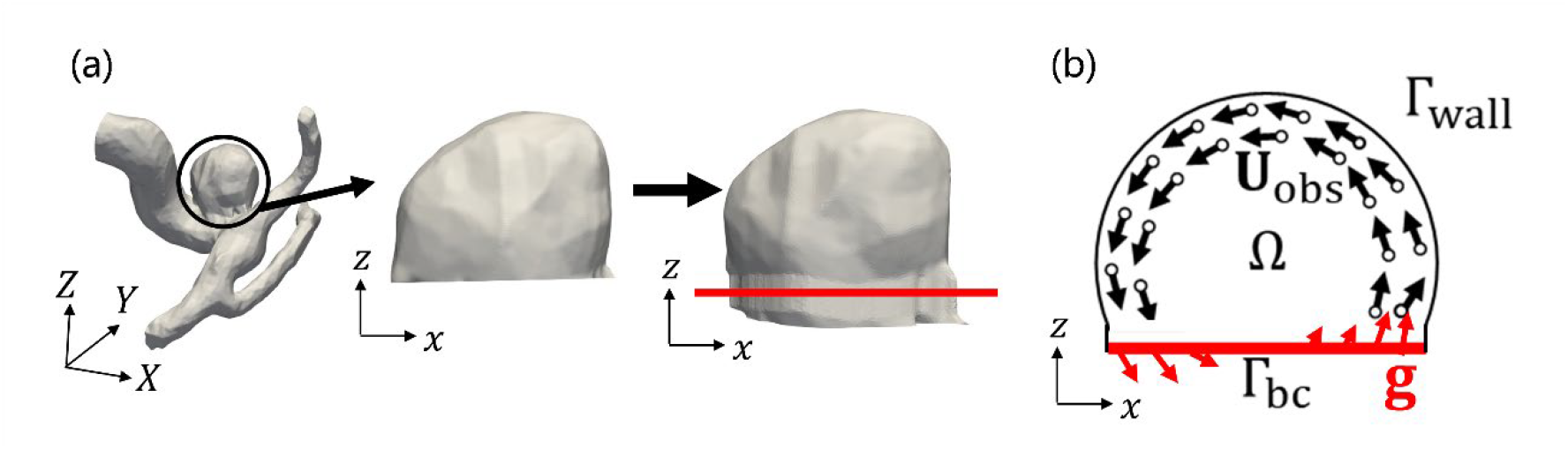
(a) Aneurysm geometry for the data assimilation (DA). (b) Solution domain of the inverse problem to obtain the observed velocity U_obs_(x, *t*) and an inlet/outlet velocity g in the space x ∈ Ω enclosed by the fixed wall Γ_wall_ and a cross-section through the aneurysm neck Γ_bc_.

By applying the Lagrange multiplier technique, the minimization problem (1) can be rewritten as

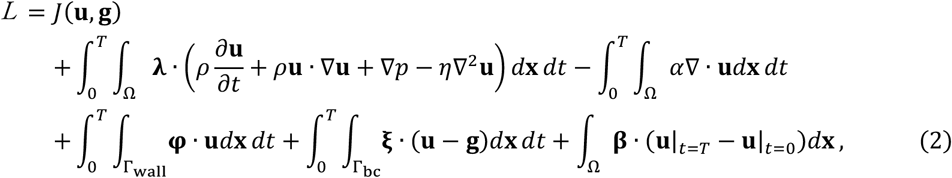

where **λ**(**x**, *t*), *α*(**x**, *t*), **φ**(**x**, *t*), **ξ**(**x**, *t*) and **β**(**x**) are the Lagrange multipliers. The stationary conditions result in sets of partial differential equations (PDEs) for the original (state) system

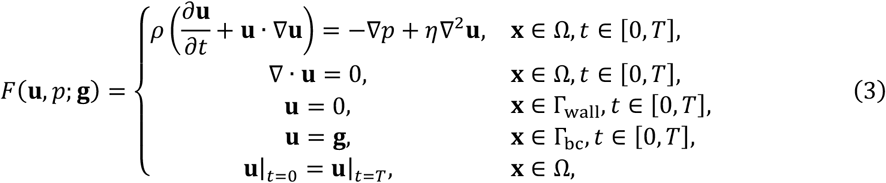

and the adjoint system

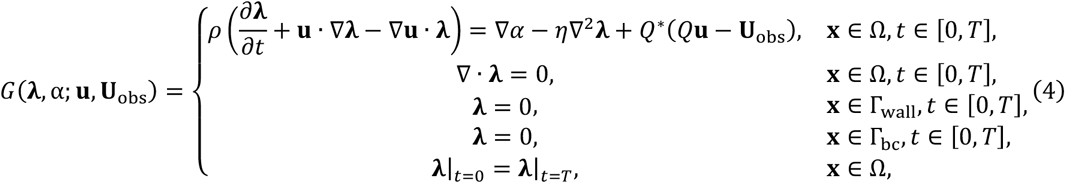

where *Q*^*\**^ is the inverse operator mapping the observation to the model field. Analogously, the functional derivative of *J* in terms of **g** is given by

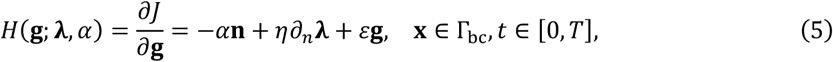

where *∂*_*n*_ = **n** · ∇ is the normal derivative on the surface Γ_bc_. More details of the derivation are provided in Appendix B.

Note that the equality *H* = 0 gives first-order optimality conditions of the optimization problem, and in general the above nonlinear systems are iteratively solved for the design variable **g** so that *H* → 0.

#### 2.2.2. Optimization calculation

A gradient descent method was used to solve the optimization problem. From Eq. (5), the evolutional equation for **g** is then given by

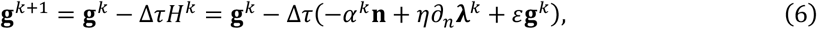

where *k* enumerates the optimization step and Δ*τ* is the hyperparameter in the gradient descent to control corrections in the optimization interval [*k, k* + 1]. The model and adjoint variables are obtained by solving systems (3) and (4) as *F*(**u**^*k*^, *p*^*k*^; **g**^*k*^) and *G*(***λ***^*k*^, *α*^*k*^; **u**^*k*^, **U**_obs_), where the pulsatile periodicity in flow should be sufficiently satisfied. System (4) was solved numerically in a time-reversed manner from *t* = *T* to *t* = 0.

#### 2.2.3. Model order reduction

Focusing on the periodicity of the problem, model order reduction was implemented to express temporal changes in variables, including the design variable, in terms of Fourier series expansions. Let us denote the Fourier series expansion of **g** by

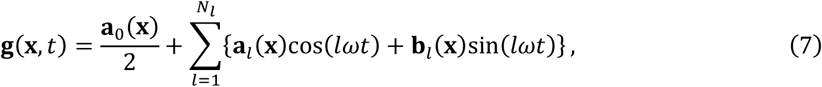

where *N*_*l*_ is the maximum expansion order, *ω* = 2*π*/*T* is the angular frequency, and **w**_*l*_ (**x**) and **b**_*l*_ (**x**) are the Fourier coefficient distributions on Γ. Denoting 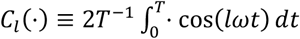 and 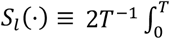, the mappings to evaluate coefficients in the Fourier cosine and sine series in Eq. (7), namely **w**_*l*_ (**x**) = *C*_*l*_ **g**(**x**, *t*) and **b**_*l*_ (**x**) = *S*_*l*_ **g**(**x**, *t*), can be rewritten as

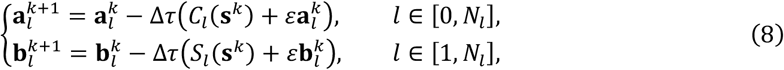

where **b**^*k*^ = −*α*^*k*^**n** + *η∂*_*n*_**λ**^*k*^. Thus, once the Fourier coefficients for the term **b**^*k*^ are evaluated, **w**_*l*_ and **b**_*l*_ can be updated using Eq. (8). In this study, *C*_*l*_ (**b**^*k*^) and *S*_*l*_ (**b**^*k*^) were calculated using trapezoidal numerical integration at the same time as solving the adjoint system (4).

The present method of model order reduction has three potential advantages. First, it mitigates an ill-posed inverse problem resulting from large degrees of freedom when directly updating **g**, which is discretely introduced in every time step of the fluid simulations. Second, the continuous profiles of the Fourier series smooth temporal changes. Third, the method dramatically reduces the memory required to store temporal changes in **g** and **u** when solving the adjoint system (4).

Note that to satisfy the flowrate condition 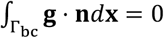, the Fourier coefficients are modified by 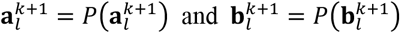, where the modification function *P*(**d**) is simply defined as a shift operator with its spatial average in the normal component 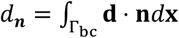 given by *P*(**d**) **= d −** *d*_*n*_**n**.

#### 2.2.4. Mapping operators between modelled and observed data

The mapping operator *Q* was defined as

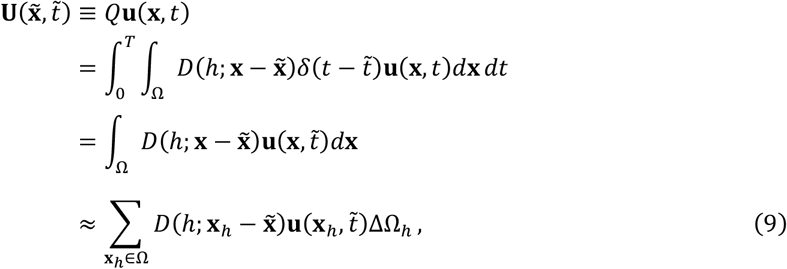

where the kernel *D* is a function of a parameter *h, δ* is the Dirac delta function, and ΔΩ_*h*_ is the size of small control domain **x**_*h*_ ∈ Ω (in general, ΔΩ_*h*_ = *h*^3^). Note that the observation position is not guaranteed to equal that of the model, and thus smoothing is required by applying the kernel at the resolution of the CFD mesh. Since a Cartesian mesh was used to solve the fluid equations, as described later, a three-dimensional smooth Dirac delta function was used for *D* [19,12]. Analogously, the inverse mapping operator *Q*^*\**^ was defined as

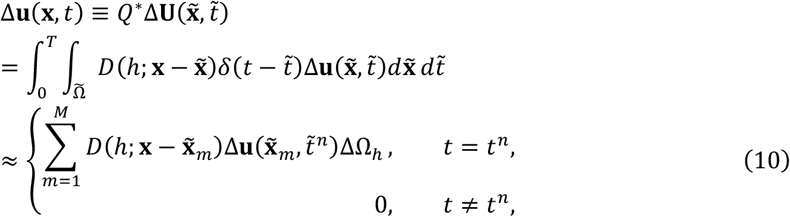

where 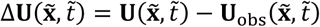 is the velocity mismatch between the model and observation, and 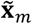 and 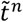 are the discrete position and time, respectively, associated with the observation data, namely 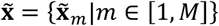 and 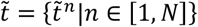. Here, *M* and *N* are the number of observation data for space and time, respectively. The first term of the objective function *J* in Eq. (1) was approximated by

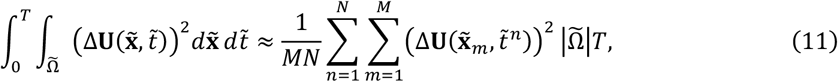

where 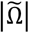 is the observation domain size.

### 2.3. CFD solver

The original and adjoint systems in Eqs. (3) and (4), respectively, were solved using a Cartesian mesh-based CFD solver [12,20] based on the boundary data immersion method [21]. This method solves a unified PDE model for fluid and solid dynamics using a smooth characteristic (or density) function, so a simple fixed-mesh system such as the Cartesian coordinate system can be adopted. Analogous to the existing solver [12,20], a pressure projection method was used to couple the velocity and pressure (and the related adjoint variables) on a Cartesian mesh.

The following material parameters were used: *ρ* = 1050 kg/m^3^, *η* = 0.0035 Pa. s [12, 22, 23]. The dimensions of the mesh elements were set to Δ*x* = Δ*y* = Δ*z* = 0.2 mm, and the time interval was set to Δ*t* = 0.05 ms, yielding a Courant number of approximately 0.1 for sample B and 0.2 for samples A and C with respect to their corresponding maximum velocities. Simulations were executed on single CPU (A64FX, Fujitsu, Japan) of either the Fugaku (RIKEN Center for Computational Science, Kobe, Japan) or Flow Type I (Information Technology Center, Nagoya University, Japan) supercomputers.

### 2.4. Analysis cases

#### 2.4.1. Forward simulation using main vessel branches and the observed inlet flow condition

To validate the present DA, CFD simulations using main vessel branches and observed inlet flow condition from the 4D flow MRI at VENC120 (termed “forward simulation”) were performed and compared with the DA solutions. Here, the validation was performed for sample C. In the simulations, the aneurysm and the main vessel branches were included in the analysis domain. A Dirichlet boundary condition for the velocity was imposed on a cross section of an internal carotid artery inlet and prescribed a uniform spatial profile and a Fourier series-based temporal profile evaluated from 4D flow MRI data at VENC120 around the inlet region. The outflow condition, −*p***n** + *η∂*_*n*_**u** = 0, is imposed on the outlet branch of the main vessels. The pulsatile period *T* would be ideally set to be the same as the heart period (∼ 1 s); however, no obvious change for flow unsteady properties were seen for above a certain *T*. Therefore, to reduce computational time, the pulsatile period was set to *T* = 480 ms in this study. The simulations were performed for five pulsatile periods, and the result at the final period was used for evaluation.

The observed velocities in the aneurysm region were smaller than those in the main branches and had peak magnitudes of less than 40 cm/s. Moreover, the velocity fluctuations resulting from VENC40 were also lower than those from VENC120 (Appendix C). This implies that the intra-aneurysmal flow data at VENC40 are more reliable than those at VENC120 in terms of the signal-to-noise (S/N) ratio. Therefore, the inlet velocities used in the forward simulations were adjusted so that the velocity in the aneurysm was close to that measured using 4D flow MRI at VENC40.

#### 2.4.2. Comparison of DA using synthetic data and 4D flow MRI data

Eqs. (3) and (4) were evaluated using two pulsatile times that almost satisfied the flow periodicity, and the solutions at the second time were used for evaluation of the Fourier coefficients in Eq. (8). From preliminary trials, *N*_*l*_ = 5 was set for the Fourier-series expansion, Δ*τ* = 2000 was set in the gradient descent. To avoid local optima, we ran several times of DAs with random in-plane orientations set as initial conditions for the control variable (boundary velocity) **g**, and used the optimal solution as the solution. In this regard, the Tikhonov parameter was set so that the DA solution was obtained stably. The number of iterations of the optimization was set to 300. The parameter *h* for the kernel *D* mapping the velocity between the model and observation was set to *h* = Δ*x* so that the kernel was smoothly distributed across neighboring mesh elements, and observation data at positions two or more mesh elements away from the wall and inlet/outlet boundaries was used.

For validation, DA was performed in the aneurysm region using synthetic data created from a forward simulation by down-sampling its spatiotemporal resolution to match that of the 4D flow MRI data. The numerical results of the forward simulation were then used as the ground truth.

For practical applications, DAs were conducted using 4D flow MRI data acquired at VENC40 for three patients.

### 2.5. Metrics for velocity mismatch

Using the L2 norms

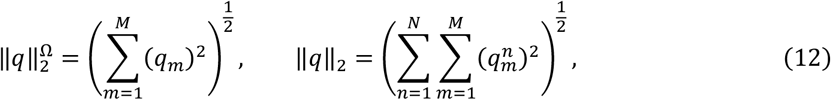

for spatial and spatiotemporal quantities, where *q*_*m*_ = *q*(**x**_*m*_)|_*t*_ and 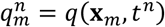, normalized metrics for the velocity mismatch between the model and observation (*E*_*u*_) were defined in space and time:

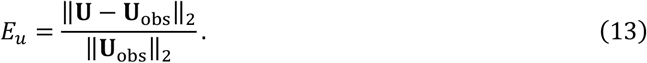

Other metrics for the velocity mismatch between the model and observation were also introduced with respect to the vector (*R*_*u*_), magnitude (*R*_|*u*|_), and angle (*R*_∠*u*_):

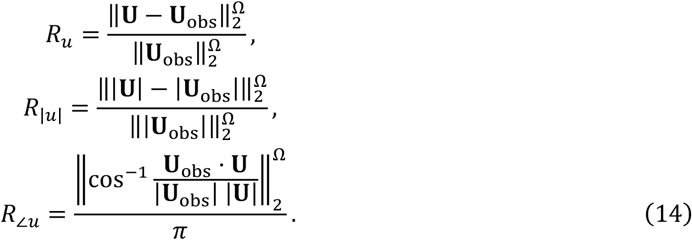

## 3. Results

### 3.1. Validation using synthetic data

Fig. 3 shows estimated results of the DA using synthetic data for sample C and the ground truth obtained from the forward simulation. The DA well captured the velocity distributions in two cross sections. The velocity mismatch is converged in optimization process and the instantaneous value in each frame fell within the 4%–7% range. In the longitudinal cross section, some discrepancies between DA and the ground truth were observed around the aneurysm neck. The forward simulation as the ground truth resulted in high velocities at the aneurysm neck and a corresponding inlet jet along the side wall. However, the DA did not adequately capture these features. This might explain the maximum mismatch at frame 11, at which time the jet-like inlet flow increased from a low-flow state as the systolic phase was approached.

**FIGURE 3.**
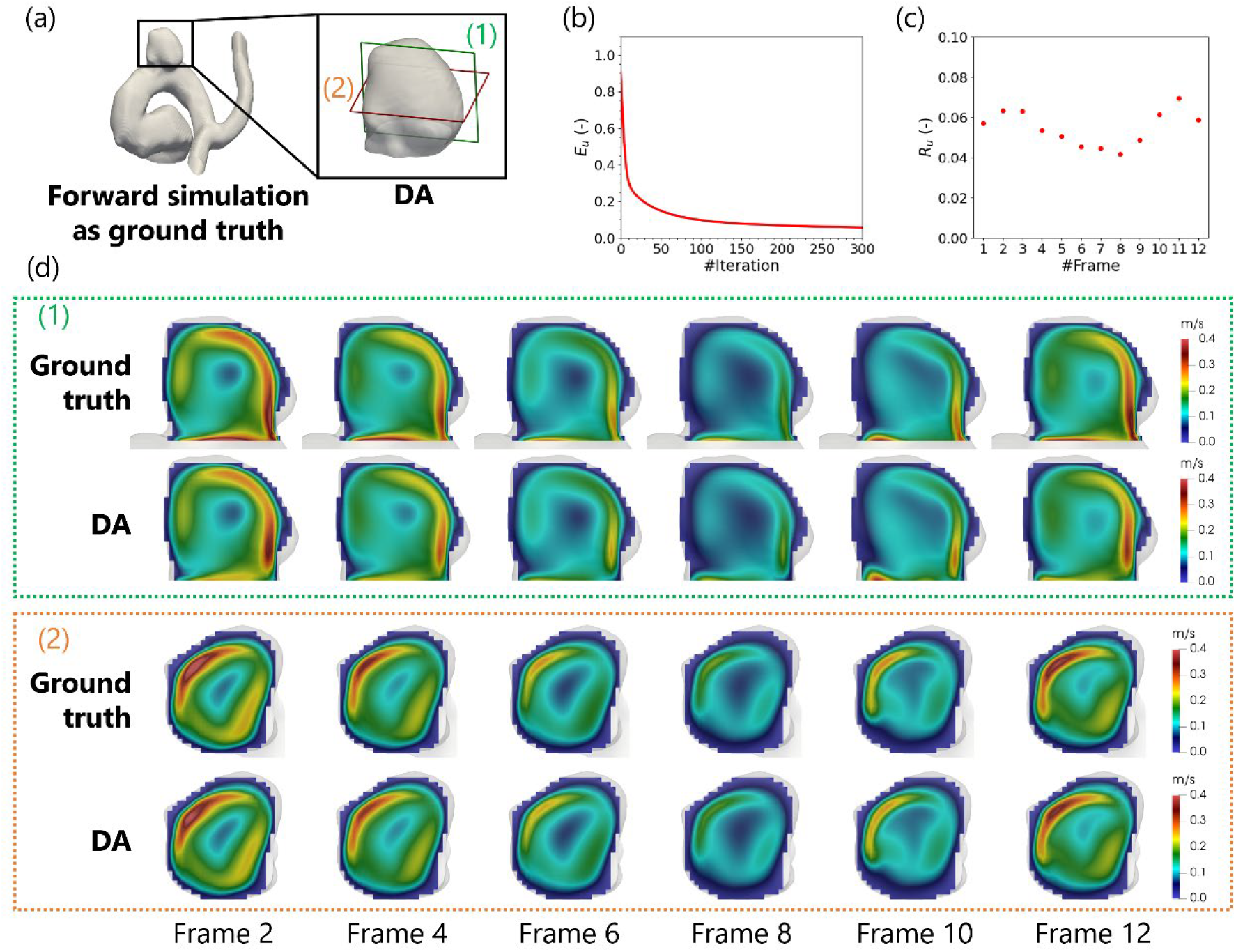
(a) Vessel geometries for the forward simulation as ground truth and DA for sample C. (b) Convergence behavior of velocity mismatch between the ground truth and DA in optimization process. (c) Velocity mismatch between the ground truth and DA across 12 frames. (d) Velocity distributions in a longitudinal cross section (1) and transverse cross section (2) at frames, 2, 4, 6, 8, 10 and 12. The systolic and diastolic phases occur at approximately frame 2 and frame 8, respectively.

Fig. 4 compares the velocity in the ground truth with that obtained from DA, both of which are plotted at the observation resolution. Excellent agreement was achieved for velocity vectors in the systolic phase. Although the Bland–Altman plot shows there was some additive bias (mean) and fluctuation (standard deviation) in mismatches of the velocity magnitude, the sizes of these mismatches were much smaller than the velocity values themselves.

**FIGURE 4.**
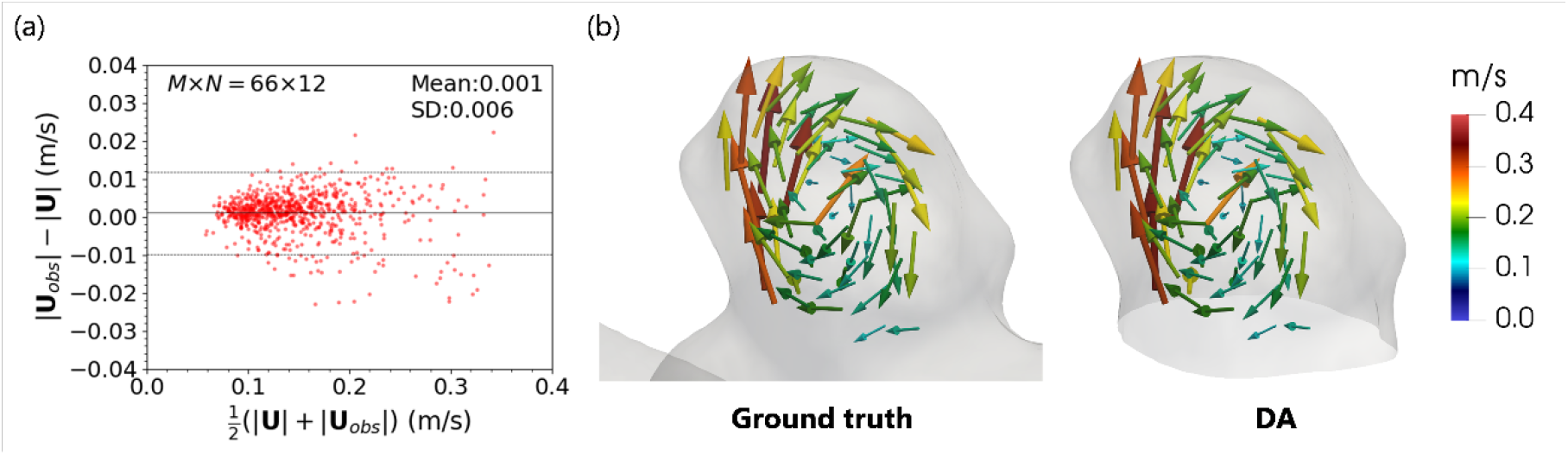
Comparison of the velocity in the ground truth with that obtained from DA using the synthetic data for sample C. (left) Bland–Altman analysis for the velocity magnitude for the DA |U| and synthetic data |U_obs_| in all frames, where the mean and standard deviation (SD) are 0.001 m/s and 0.006 m/s, respectively. (right) Velocity vectors in the systolic phase. The solid and dashed lines denote the mean and mean ± 1.96 SD, respectively.

### 3.2. Comparisons with 4D flow MRI data

The intra-aneurysmal velocity vectors obtained from 4D flow MRI, the proposed DA, and forward simulation at systolic phases for three samples are shown in Fig. 5. It should be noted that the optimal solutions for various initial boundary conditions are presented here, but unlike the validation using synthetic data, the converged solution using the actual dataset was not largely dependent on these initial conditions. Although both the DA and forward simulation could capture the overall behavior of the flow fields obtained from 4D flow MRI, there were several large discrepancies in the velocities between samples B and C. The mismatches (Eq. (12)) between the velocity obtained from 4D flow MRI and those obtained from the forward simulation and DA are summarized in Table 1. For all three samples, the velocity mismatches between the 4D flow MRI and DAs were 37%–44% lower than those between the 4D flow MRI and the forward simulations. However, the velocity mismatches between the DA and 4D flow MRI analyses for samples B and C were approximately twice as large as the corresponding mismatch for sample A. The velocity magnitudes (i.e., the spatiotemporally averaged velocities using the L2 norm) for the 4D flow MRI were 0.132, 0.095, and 0.129 m/s for samples A, B, and C, respectively. Notably, the size of these values was in the order A > C > B, which followed the opposite trend of the velocity mismatches (A < C < B).

**Table 1.** Mismatches between the velocity obtained from 4D flow MRI and the velocities obtained from the forward simulation and data assimilation (DA) analysis for samples A, B, and C.

|  | Sample A | Sample B | Sample C |
| --- | --- | --- | --- |
| Forward simulation | 0.79 | 1.71 | 1.51 |
| DA | 0.35 | 0.63 | 0.60 |

**FIGURE 5.**
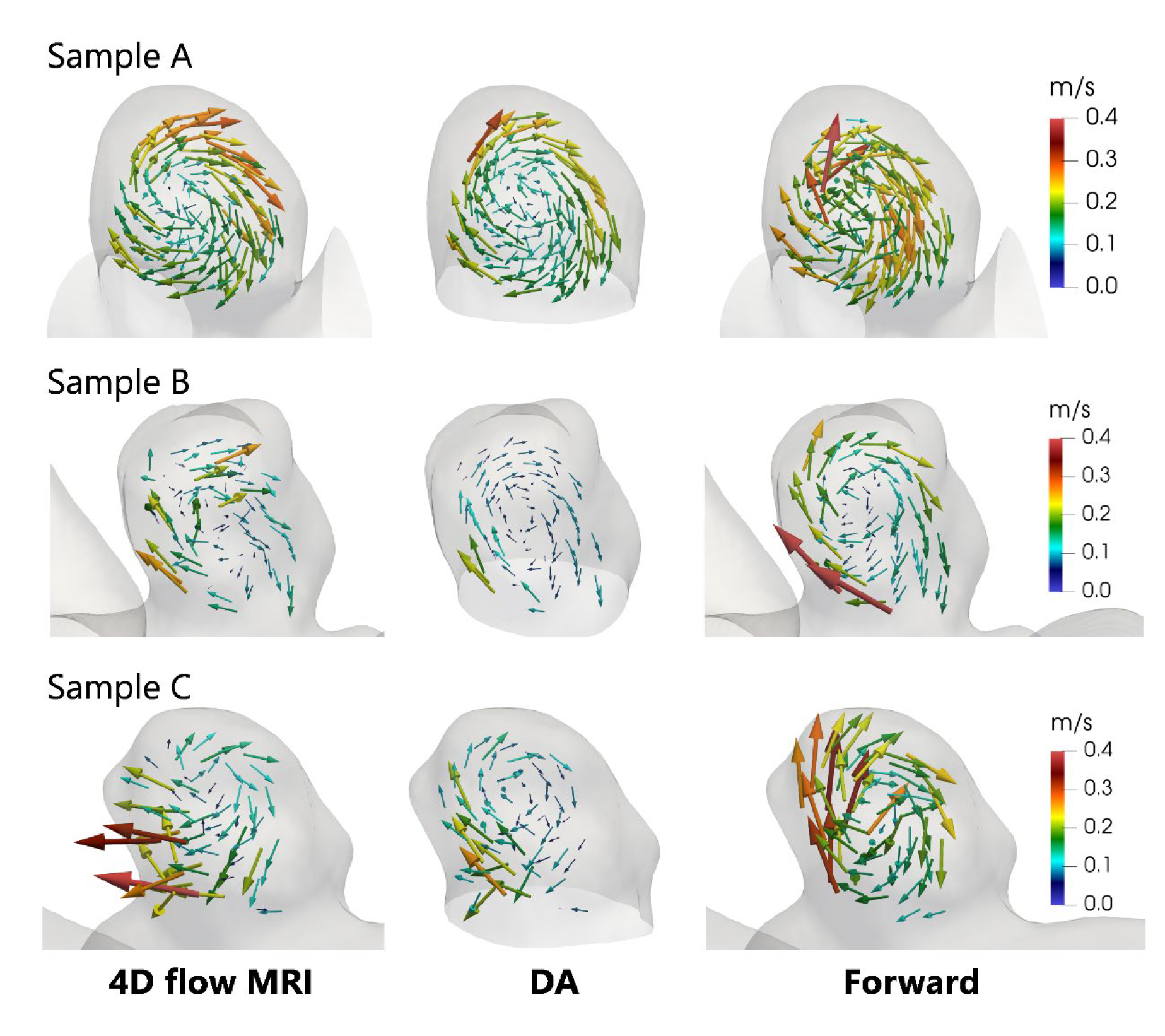
Comparison of velocity vectors in cerebral aneurysms during the systolic phase, obtained from the proposed DA, 4D flow MRI, and forward simulations.

Bland–Altman plots of the agreement between the velocity magnitude obtained from 4D flow MRI and velocities obtained from the DA and simulation are shown in Fig. 6. For all samples, velocity differences with the 4D flow MRI were much smaller for the DA than for the forward simulation. Moreover, variation in the velocity differences among the samples was smaller for the DA. There is a tendency that velocity mismatch shifts in one direction as the velocity magnitude increases in the DA for samples B and C.

**FIGURE 6.**
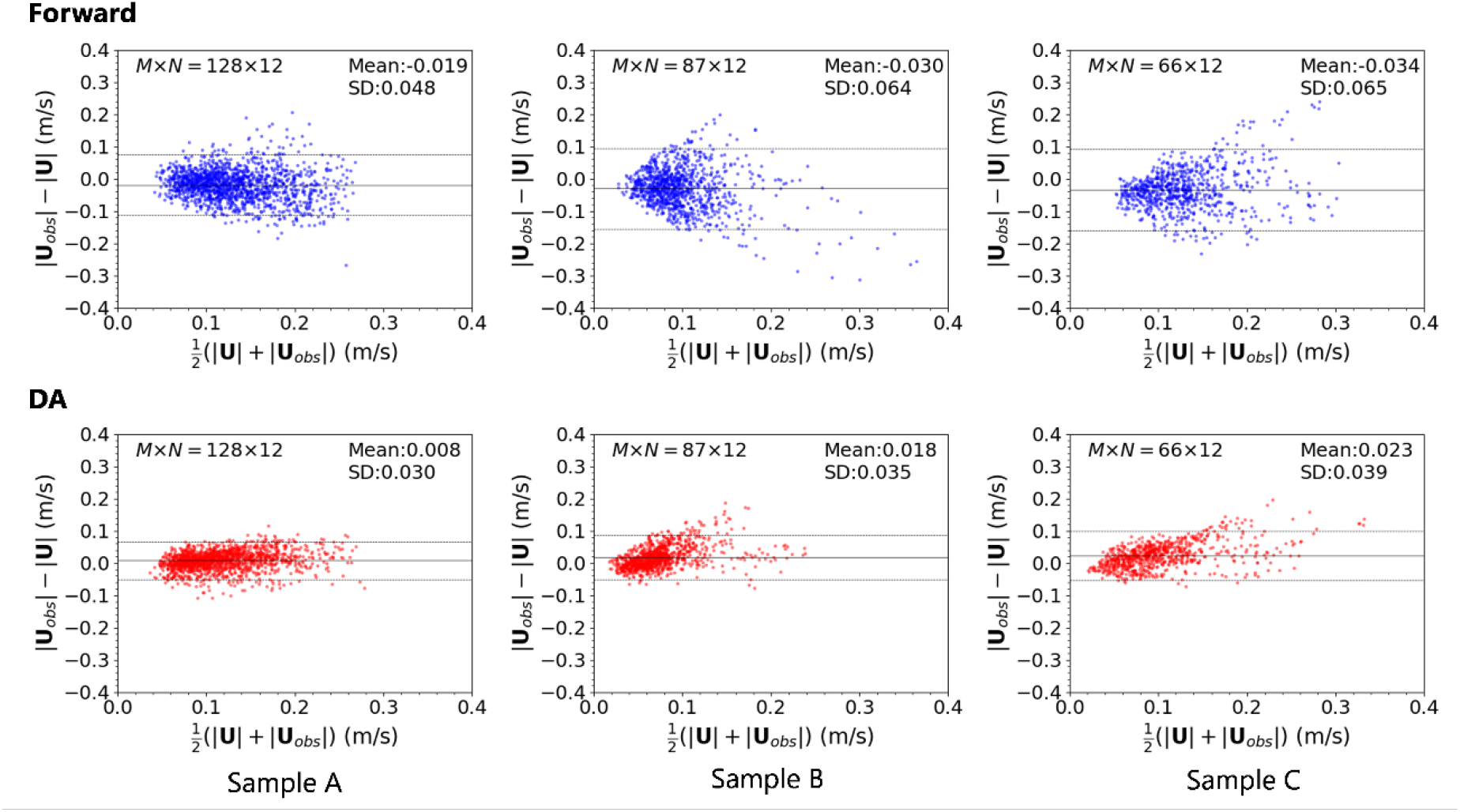
Bland–Altman plots of the agreement between the velocity magnitude obtained from 4D flow MRI and the magnitudes obtained for samples A, B, and C from the forward simulation (top) and DA (bottom). The solid and dashed lines denote the mean and mean ± 1.96 SD, respectively.

To further investigate velocity mismatch between the DA and 4D flow MRI, metrics for vector, magnitude, and angle in Eq. (14) were evaluated (Fig. 7). The metric for the magnitude was always lower than that for the vector, and the angle metric was approximately 0.1–0.3 (i.e., 10°–54°). Again, the size of the metric mismatch for samples A–C was in the order A < C < B, which followed the opposite trend of the velocity magnitudes (A > C > B).

**FIGURE 7.**
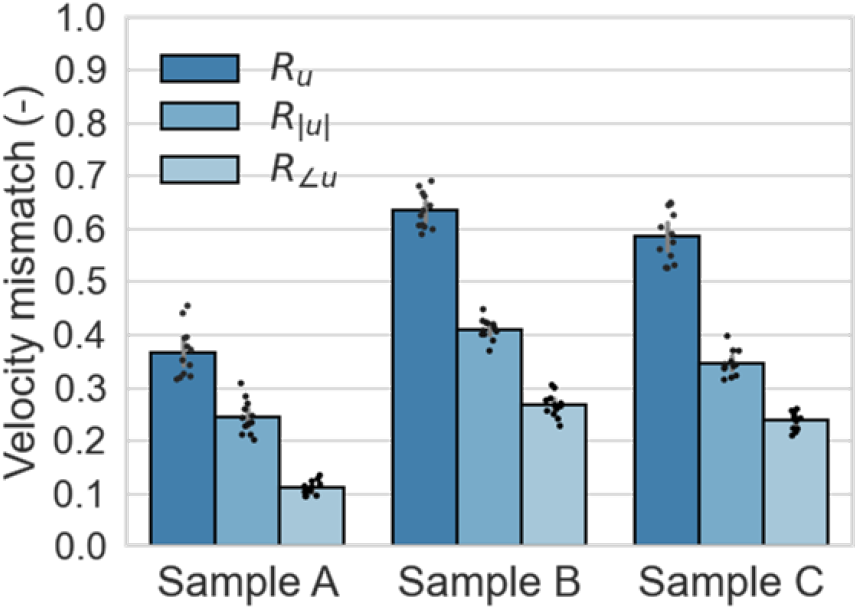
Velocity mismatches between the 4D flow MRI and DAs of the vector, magnitude, and angle metrics for samples A, B, and C.

### 3.3. Effects of VENC values on the DA

To investigate the influence of data noise on the velocity estimation, DA using VENC120 data was performed for sample A. The velocity mismatches for both VENC40 and VENC120 are shown in Fig. 8. The velocity mismatch at VENC40 was clearly much smaller than that at VENC120.

**FIGURE 8.**
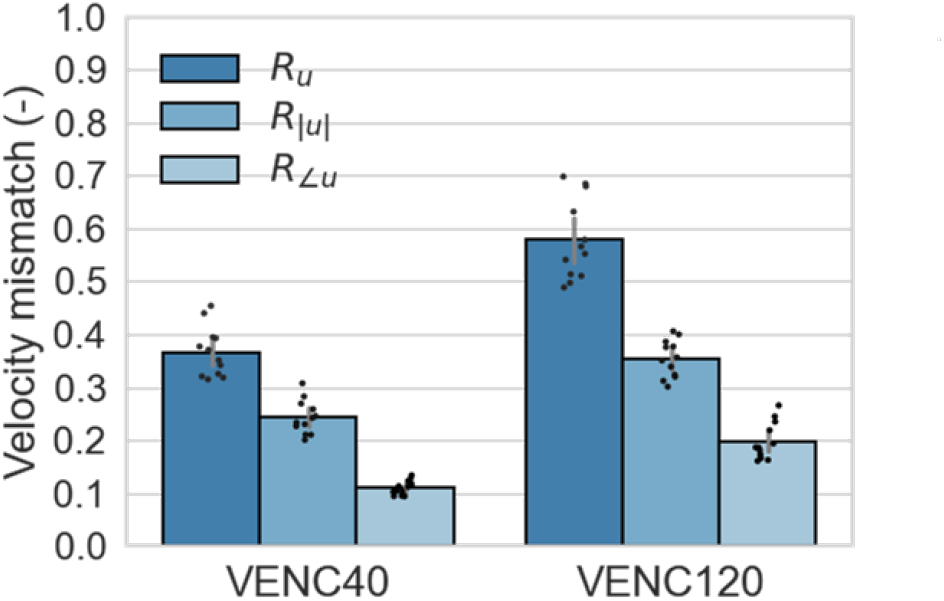
Velocity mismatches between the 4D flow MRI and DAs of the vector, magnitude, and angle metrics for sample A, using 4D flow MRI velocity encoding (VENC) at 0.4 m/s (VENC40) and 1.2 m/s (VENC120).

### 3.4. Evaluation of flow patterns and hemodynamic factors

We investigated flow streamlines in the DA and forward simulations, obtained from detailed flow profiles at CFD resolution (Fig. 9). Overall circulation patterns within the aneurysm appear to be similar in the DA and forward simulations, with high velocity regions near the aneurysm sidewall and a main vortex core being generated. However, the jet-like patterns at the aneurysm entrance show large discrepancies in samples B and C. Fig. 9 also shows the streamlines of the main vessel in the forward simulation showing how the flow enters the aneurysm, with seed points of the streamlines randomly set at the aneurysm entrance. A portion of the separated flow from the main vessel enters the aneurysm, showing a helical flow structure, which is sensitive to the main vessel geometry, the presence of branch vessels, and the boundary conditions.

**FIGURE 9.**
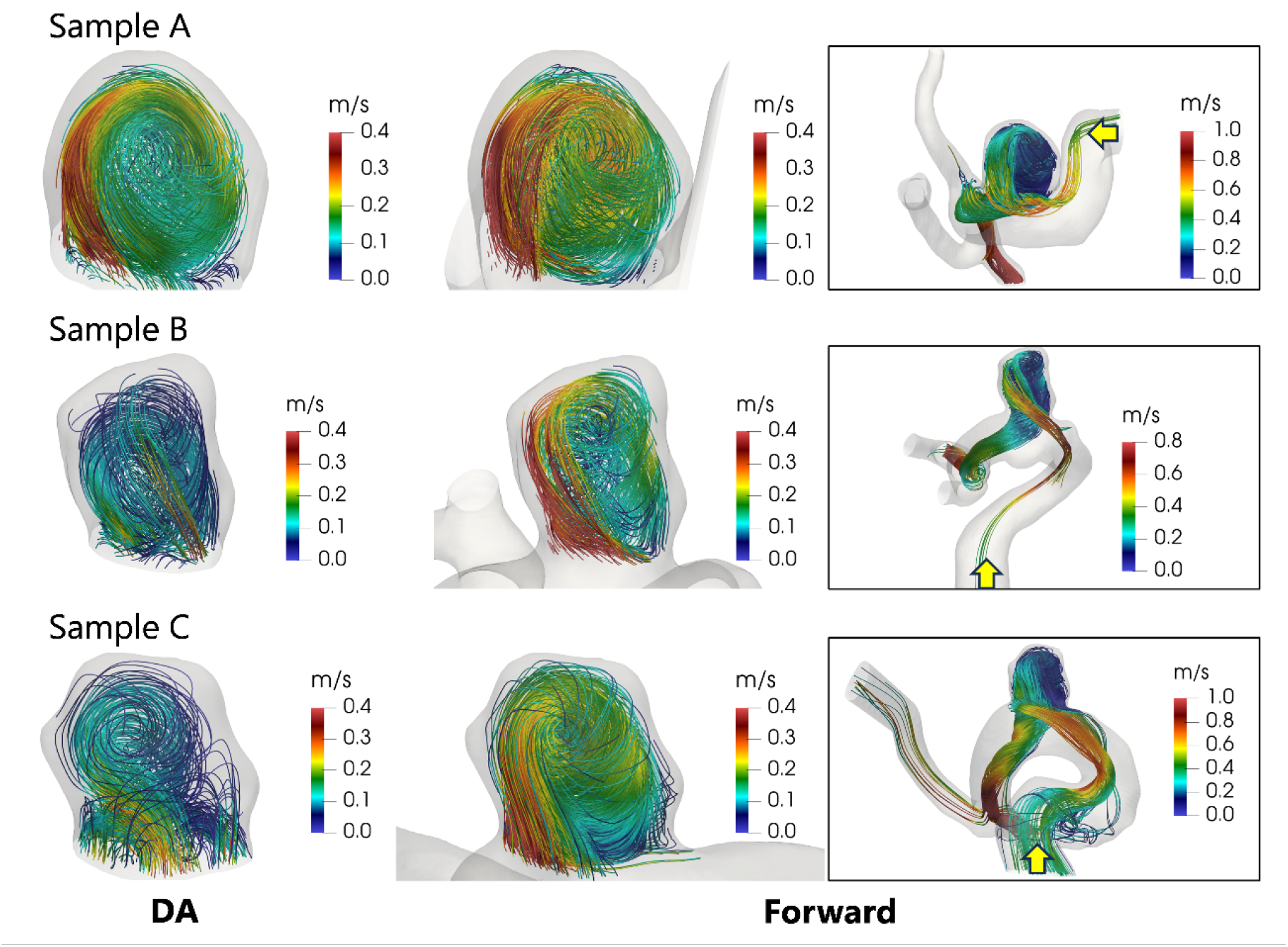
Comparison of flow streamlines during the systolic phase, obtained from the proposed DA and forward simulation. The yellow arrow in the right figures denotes the direction of the main blood flow.

High-resolution velocity fields allow the assessment of wall shear stress (WSS) and therefore time-averaged wall shear stress (TAWSS) and oscillatory shear index (OSI), which are widely used as hemodynamic factors:

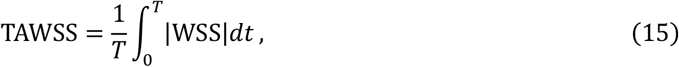

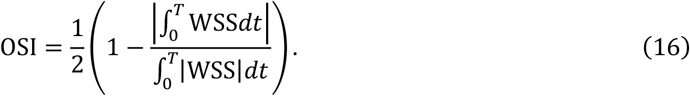

In this study, a Cartesian-based method [12] is applied to calculate the WSS. The TAWSS, OSI and pressure field during systolic phase are shown in Fig. 10. High OSI areas tend to appear around low TAWSS, and the distribution and quantitative values, such as low pressure around the central core of the aneurysm due to blood flow circulation, are in good agreement with existing reports [10, 24].

**FIGURE 10.**
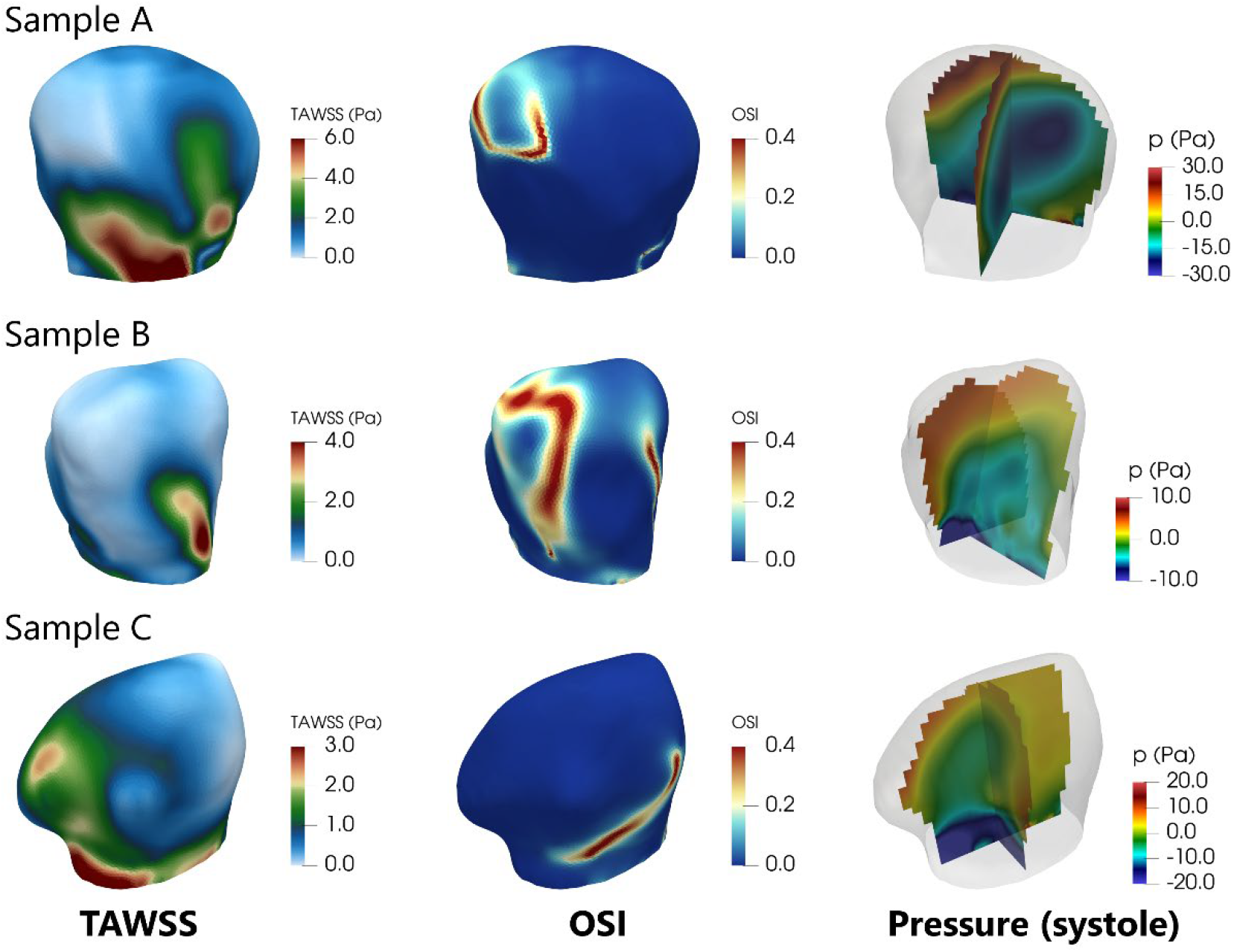
Distributions of TAWSS, OSI, and cross-sectional pressure during systole, obtained from the proposed DA.

## 4. Discussion

A novel numerical approach to quantify patient-specific intra-aneurysmal flows was developed using a 4D-Var DA. The proposed DA approach focuses on an intra-aneurysmal analysis domain and inversely estimates a spatiotemporal profile of boundary velocity at the aneurysm neck. This could dramatically reduce the calculation time in forward simulations for the original and adjoint systems in every optimization step. Moreover, the proposed approach bypasses an uncertainty of an image processing-based extraction procedure for main vessels, which is sensitive to the CFD [24,25,26]. Using model order reduction to describe temporal changes in variables offers further advantages for avoiding an ill-posedness in an inverse problem and reducing the corresponding memory requirement. Although a similar idea to parametrize the spatiotemporal profile of the inlet velocity with a few control parameters was previously proposed [17], the method could not straightforwardly apply to our approach because it is restricted to simple velocity profiles in both space and time. For these reasons, the proposed DA approach is anticipated to become a practical strategy for estimating patient-specific inter-aneurysmal flows.

Using the synthetic data obtained from the forward simulation in an aneurysm and the main vessel branches with a boundary condition imposed on vessel geometry, the present DA could reasonably reproduce the original (ground truth) flow field. The spatially averaged velocity mismatch was within approximately 5% in the systolic phase (until frame 7) and 7% between the end of diastole (frame 8) and the beginning of the next systole (frame 10). During these frames, a jet-like inflow from the aneurysm neck resulted in high flow instability and sudden changes in the flow magnitude and direction, which would lead to a relatively large error. The objective function for the velocity mismatch did not converge to zero even in the DA using the synthetic data. In this validation, the inlet/outlet boundary of the aneurysm was vertically extended from the neck to stabilize the numerical simulations, and this change in aneurysm shape may have caused a mismatch with the ground truth. This is consistent with the observation that the estimation accuracy of the present DA was lower around the aneurysm neck than inside the aneurysm. Notably, although the above scenario may have occurred, reasonable flow estimations inside the aneurysm were nevertheless obtained.

In image-based blood flow simulations, hemodynamic factors inside an aneurysm are dependent on inflow boundary conditions [10]; nevertheless, setting complete spatiotemporal profiles on the inlet of main vessel is difficult owing to resolution and noise limitations in 4D flow MRI. Moreover, the flow field inside the aneurysm is strongly dependent on the accuracy of shape reconstruction of main vessels because the magnitude of the intra-aneurysmal flow is much smaller than that through the main vessels. This could explain why the forward simulation using main vessels reproduced the 4D flow MRI velocity with low accuracy. In contrast, the present DA, where the spatiotemporal profiles on the inlet/outlet boundary of the aneurysm neck are automatically set by solving the optimization problem, achieved 37%–44% less velocity mismatch than the forward simulation. From distribution of streamlines, the jet-like patterns at the aneurysm entrance show some discrepancies, especially large in samples B and C. The helical flow in the main vessel is sensitive to the main vessel geometry, the presence of branch vessels, and the boundary conditions. Both the forward simulation and present DA have uncertainty in the vessel and aneurysm shapes, and thus uncertainty analyses are required to accurately evaluate the flow fields [27]. In this regard, the present DA has the advantage of focusing only on the aneurysm.

The velocity mismatch between the DA and 4D flow MRI varied among samples. The order of the relative sizes of the mismatches for samples A–C was opposite that of the relative magnitudes of the spatiotemporally averaged velocities obtained from 4D flow MRI; however, the difference in the average velocity values between samples A and C was small compared with the corresponding difference in the velocity mismatches (i.e., A ≅ C > B), whereas the velocity mismatch between sample B and C was small (i.e., A < C ≅ B). When focusing on flow fields, all samples had an organized circulating flow, but several high-velocity vectors were observed in the 4D flow MRI of samples B and C, and some of them appeared to be oriented toward the aneurysm wall. The forward simulation and proposed DA based on the inverse problem, which completely follow the incompressible Navier-Stokes equations, cannot reproduce such unphysical flows seen in the 4D flow MRI. It has been found that 4D flow MRI is affected by temporal averaging of multiple heartbeats, motion artifacts and displacement artifacts [28]. Indeed, validation studies using phantom models of intra-aneurysmal flows have shown that 4D flow MRI show different flow profiles compared to other quantification techniques such as CFD, particle image velocimetry, and particle tracking velocimetry [6,7,8,29]. In addition, it should be noted that the surface shape of the aneurysm is not reconstructed well in low-velocity regions using TOF-MRA measurement [30], and this would reduce prediction accuracy for the CFD. It is worth noting that in the DA for samples B and C, there is a tendency for systematic error such that the velocity mismatch shifts in one direction as the velocity magnitude increases. At this time, the source of the large artifact for samples B and C remains unclear, and further numerical investigation using various conditions is needed to discuss how such measurement artifacts affect the DA. Furthermore, if the observation artifacts are not secondary, comparison with different types of DA, such as the Kalman filter-based method [18] or sparse representations [32], would also be important.

In 4D flow MRI, the choice of VENC is crucial to adequately quantify flow velocities. The value should be set to cover the maximum flow velocity to avoid aliasing artifacts; however, the S/N ratio is low in the low-velocity region because of phase errors in the 4D flow MRI measurements [31]. The flow velocity inside an aneurysm is generally lower than that in the main vessels, and thus VENC40 could more accurately evaluate the intra-aneurysmal flow velocity than VENC120 because it results in a larger S/N ratio. This explains why DA using VENC40 data resulted in smaller velocity errors than DA using VENC120. Since the proposed DA approach only requires observation of the intra-aneurysmal velocity, it is possible to use a low VENC value in the 4D flow MRI measurement to improve the S/N ratio.

Due to measurement artifacts, velocity fields obtained from 4D flow MRI are not necessarily ground truth, but rather have implications as reference data reflecting patient-specific conditions. Nonetheless, such incomplete data still provides an important piece to quantify patient-specific blood flow that the DA could retrieve physically correct flows. High-resolution and high-fidelity flow fields of DA allow the assessment of hemodynamic factors such as pressure and indices related to WSS (e.g., TAWSS and OSI). In this regard, the selection of general and patient-specific inflow conditions in CFD can lead to significant differences in the magnitude and distribution of WSS [10], so an assessment that reflects patient-specific conditions using DA is believed to be useful.

## 5. Limitations

This study had several limitations. First, as mentioned above, reconstruction accuracy of the aneurysm shape limited the estimation accuracy. In this regard, a notable technique that incorporates a shape factor of the vessel wall into an optimization problem as a design variable has been developed [33], and its implementation may improve the estimation accuracy of the present DA. Also, an approach based on the Darcy equation to describe the vessel wall can be trialed [34]. Second, since this study imposes a point-wise approximation to the kernel, more reasonable approximations to estimate the values may be considered (e.g., volume integral estimation using a reasonable kernel) [17, 22]. Third, although flow fields were considered as a hemodynamic factor, several other hemodynamic factors related to wall shear stress affect the growth and rupture of aneurysms [35]. Incorporation of these indices into the DA will enhance its applicability. Fourth, although the present DA has a lower computational cost than other DA approaches, which solve the problems including main vessel branches, calculations still require nearly 1 day to sufficiently converge the objective function during optimization. In addition to this, it should be noted that precomputations for setting the initial condition of the control variable and Tikhonov parameter also take time, involving preprocessing (image segmentation and registration) for real-world datasets. More efficient gradient-based optimization techniques, such as the Broyden–Fletcher–Goldfarb–Shanno method [16], will mitigate this issue. Fifth, standardized blood viscosity has a large and unpredictable impact on hemodynamic factors in CFD simulations [23]. Uncertainty analyses or extension of the optimization problem to include blood viscosity will be needed to correctly quantify patient-specific hemodynamic features in DAs. Sixth, the aneurysm analysis regions were manually extracted. However, hemodynamic factors around the aneurysm neck play an important role in pathobiological changes, and thus careful extraction procedures are required to accurately reproduce the neck shape. More sophisticated reconstruction techniques [36] will improve the determination of the analysis domain. Seventh, since the number of samples is limited to three, further studies are required to conclude the accuracy of the proposed approach.

## 6. Conclusions

In summary, a practical strategy to estimate patient-specific intra-aneurysmal flows based on the 4D-Var DA using 4D flow MRI data was introduced and validated. The proposed DA focuses on an intra-aneurysmal analysis domain and inversely estimates a spatiotemporal profile of the boundary velocity at the aneurysm neck. This significantly reduces the calculation time in solving a constrained optimization problem in the conventional approach based on 4D-Var DA using main vessels and bypasses an uncertainty of an image processing-based extraction procedure for main vessels. The proposed DA can better reproduce intra-aneurysmal flows in vivo than conventional CFD simulations and retrieve high-resolution and fidelity flows that follows exact physical laws. This allows for a patient-specific assessment of hemodynamic factors. The proposed approach is expected to become a key tool for early, patient-specific prediction of aneurysm growth and rupture.

## Ethics statement

The study was approved by the ethics committees for human research at Shiga University of Medical Science (IRB Number: R2019-227). After explaining the aim of this study and the potential for the detection of diseases in the brain, the healthy volunteers provided written informed consent and underwent MRI examinations. The patients’ MRI data were obtained using an opt-out method after their personal information was anonymized in a linkable manner. The study was conducted according to the approved guidelines of the Declaration of Helsinki.

## CRediT authorship contribution statement

**Tsubasa Ichimura:** Writing – original draft, Writing – review and editing, Visualization, Validation, Software, Methodology, Formal analysis, Data curation. **Shigeki Yamada:** Writing – review and editing, Investigation, Data curation. **Yoshiyuki Watanabe:** Writing – review and editing, Investigation, Resources, Data curation. **Hiroto Kawano:** Writing – review and editing, Investigation, Data curation. **Satoshi Ii:** Writing – review and editing, Writing – original draft, Supervision, Validation, Software, Resources, Project administration, Methodology, Funding acquisition, Conceptualization.

## Data availability

Data will be shared in a data repository after the manuscript is accepted.

## Acknowledgements

This work was supported by the MEXT Program for Promoting Researches on the Supercomputer Fugaku (Development of human digital twins for cerebral circulation using Fugaku, JPMXP1020230118) and used computational resources of the supercomputer Fugaku provided by the RIKEN Center for Computational Science (project ID: hp230208, hp240220, hp250236). Some computations were also carried out using the supercomputer “Flow” at the Information Technology Center, Nagoya University. This work was also supported by JSPS KAKENHI Grant Number JP22H00190, JP24K02557. The authors thank Prof. Shigeo Wada and Prof. Tomohiro Otani (Osaka University) for fruitful discussion; Mr. Daiki Kurokawa, Mr. Yu Taniguchi (Osaka University), and Mr. Kengo Tanaka (Tokyo Metropolitan University) for preliminary studies; and Edanz (https://jp.edanz.com/ac) for editing a draft of this manuscript.

## Appendix A: Vessel shapes and aneurysm sizes for each of the three patients (samples)

Fig. A1 shows vessel shapes and aneurysm sizes for each of the three patients, sample A, B and C, where all the aneurysms occur at an internal carotid artery. The aneurysm size of sample A was largest among the three samples, and small bumps were seen in the aneurysms of sample B and C. The heigh (H), width (W) and depth (D) of each aneurysm were roughly measured based on orientation of each main vessel.

**FIGURE A1.**
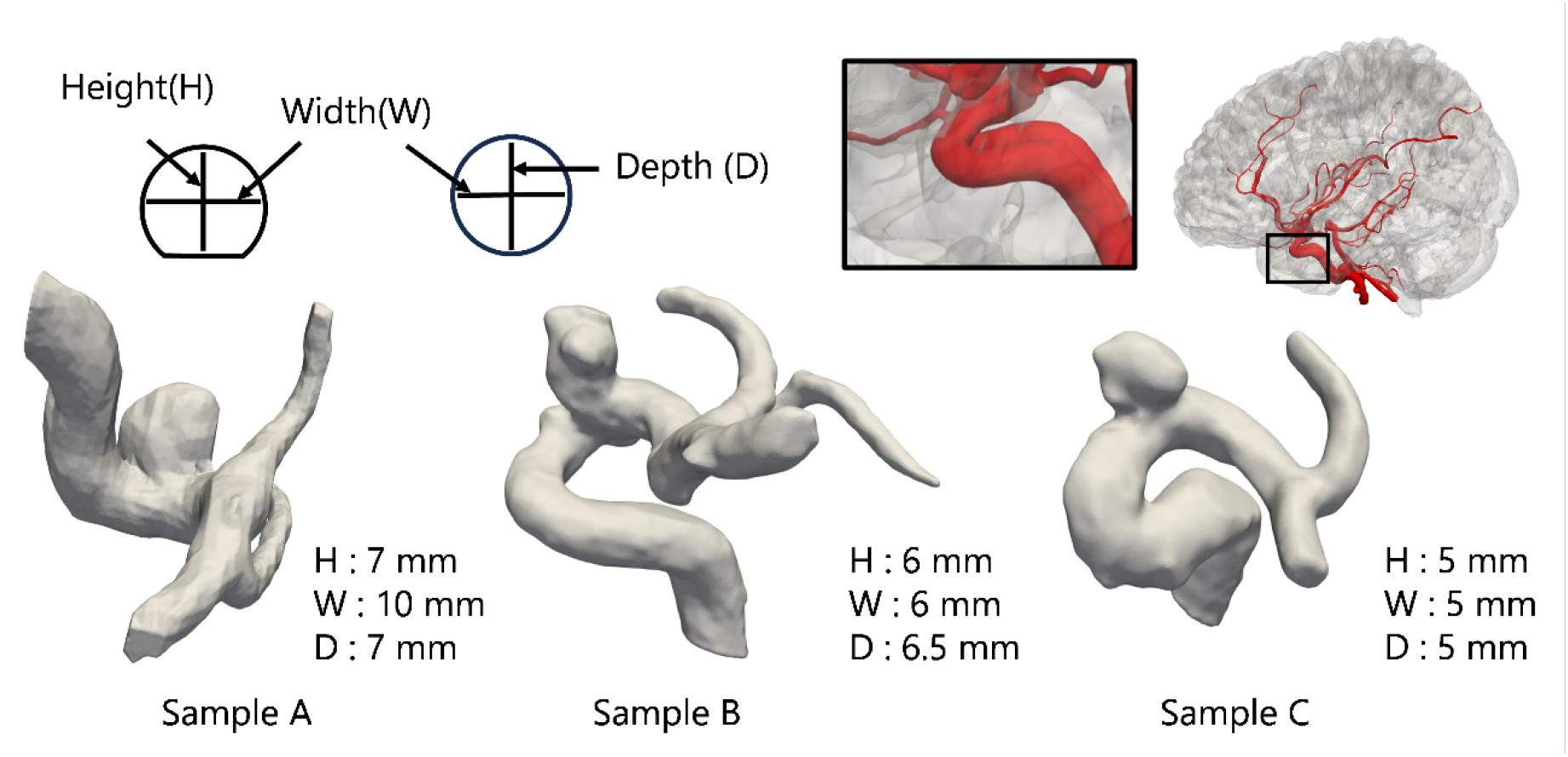
Vessel shapes and aneurysm sizes for each of the three patients (Sample A, B and C).

## Appendix B: Derivation of optimality conditions

The Lagrangian is decomposed into each component as

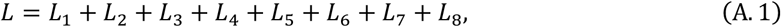

where

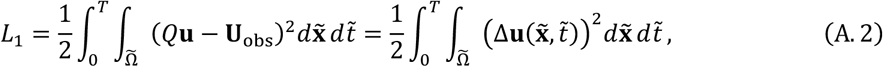

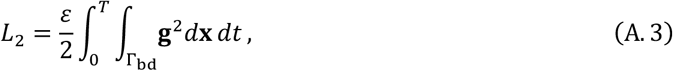

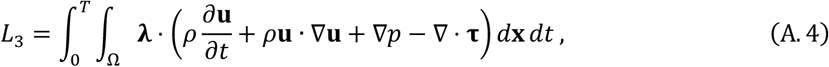

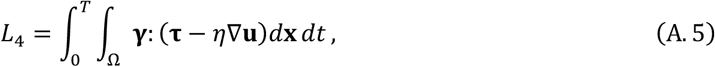

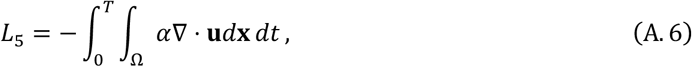

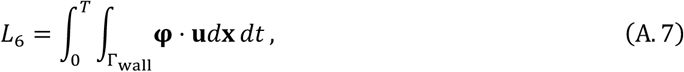

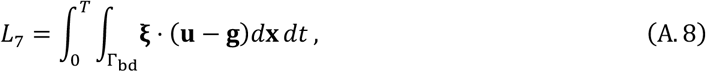

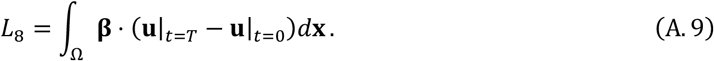

Here, the viscous term in the Navier-Stokes equations is rewritten by *η*∇^2^**u** = ∇ · **τ**, where **τ** = *η*∇**u**, and thus the term (A.5) is temporarily added to the original Lagrangian (2) with the additional Lagrange multiplier ***γ***(**x**, *t*).

Defining a variation of the Lagrangian (A.1) as 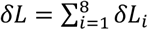, each variation *δL*_*i*_ is given as follows:

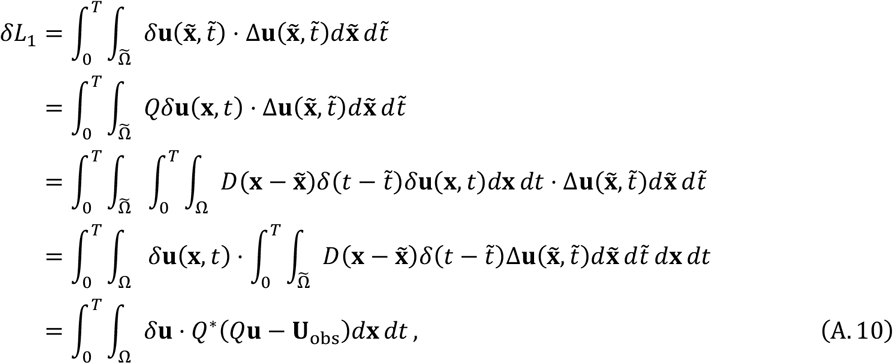

where the linear assumption is considered for *Q*^*\**^ and *Q*.

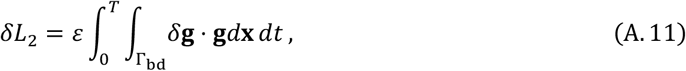

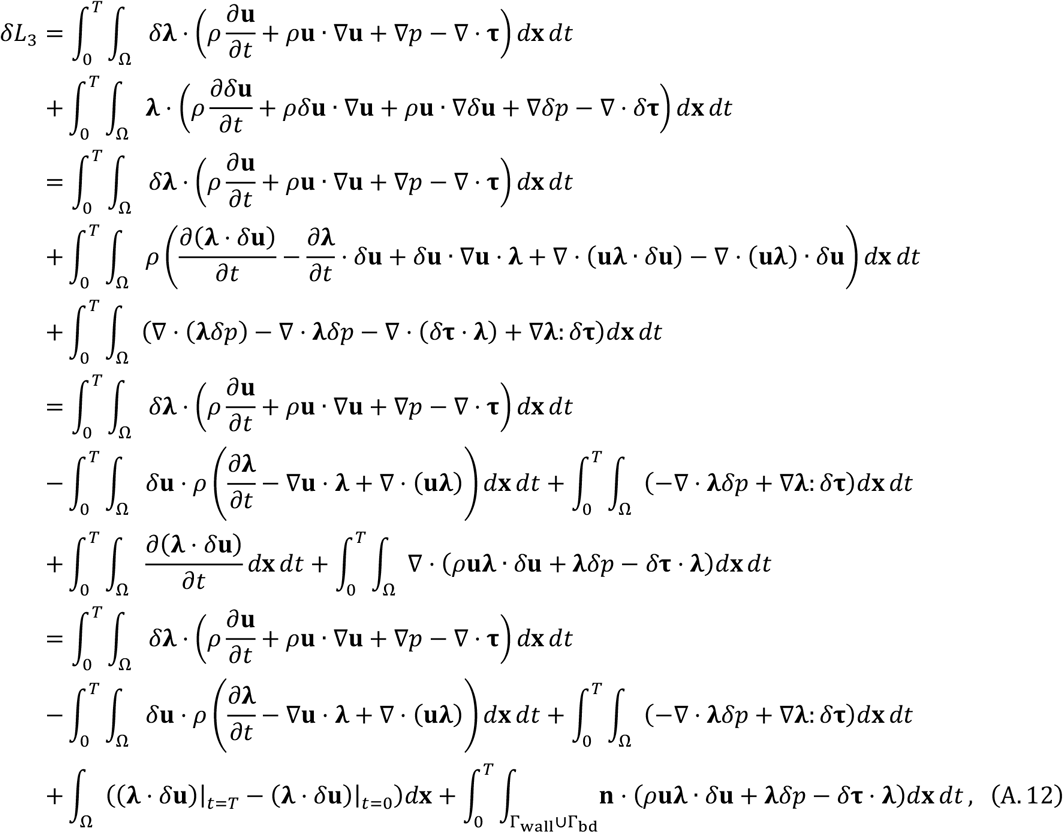

where Gauss divergence theorem is applied for the last term.

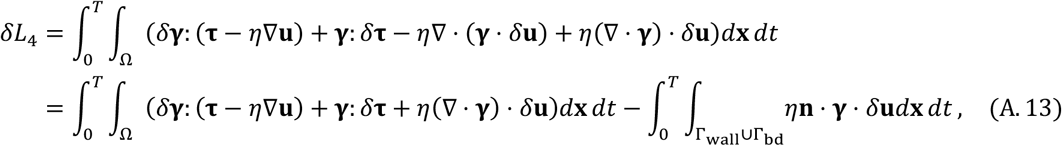

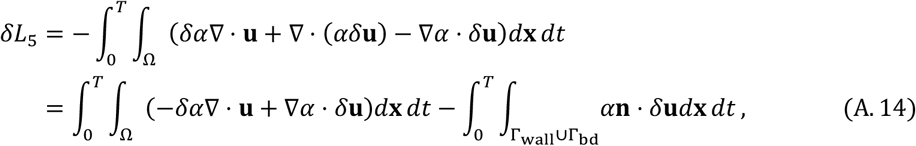

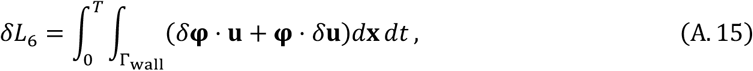

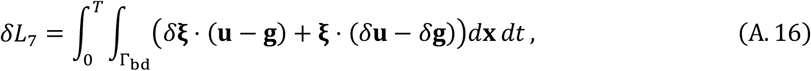

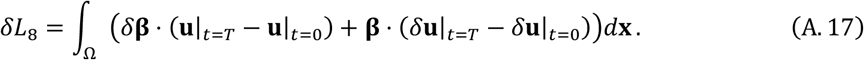

Using the above derivations, *δL* can be also written to

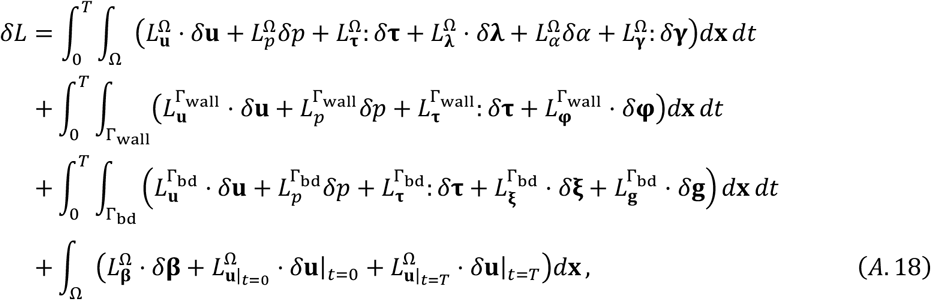

Where

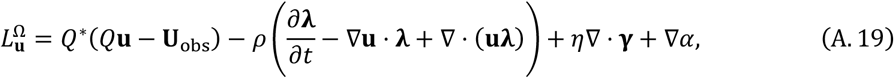

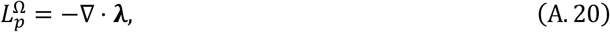

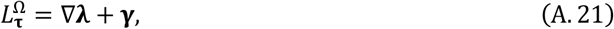

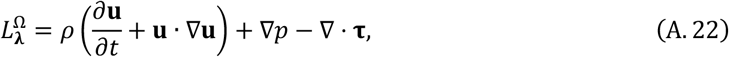

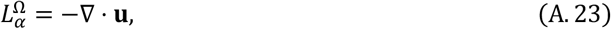

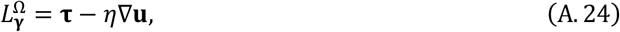

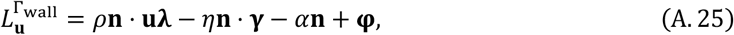

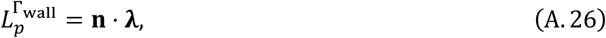

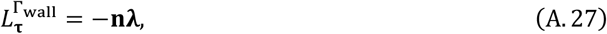

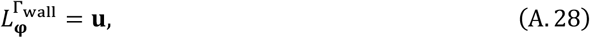

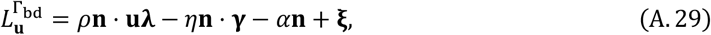

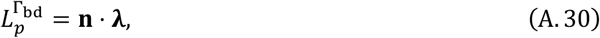

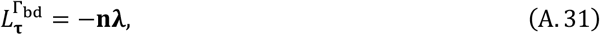

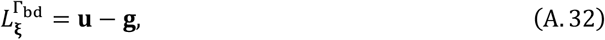

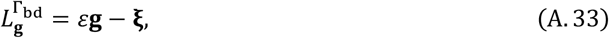

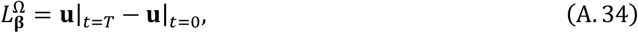

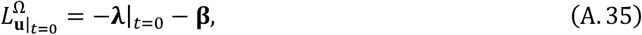

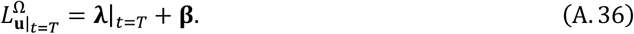

Hence, a trivial solution for *δL* = 0 is given by satisfying the following conditions:

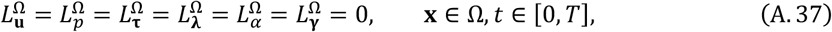

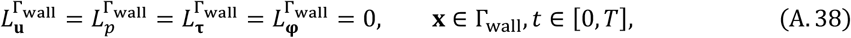

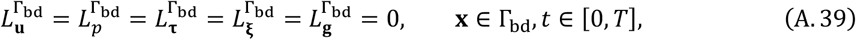

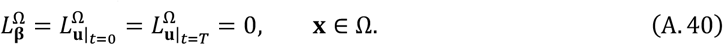

Although Eqs. (A.37)-(A.40) result in optimality conditions, a huge nonlinear system should be solved that generally causes computational costs. This study alternatively applies an iteration algorithm for satisfying the optimality conditions as described in the main text. The original and adjoint systems (3) and (4) are derived from Eqs. (A.37)-(A.40) excepting for the condition 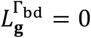, and **g** is successively updated by a gradient method using its gradient: 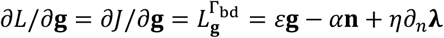.

## Appendix C: Comparison of the 4D flow MRI velocities in the aneurysm

Fig. C1 shows temporal changes of 4D flow MRI velocities at VENC40 and 120 in the aneurysm of sample A. The velocity fluctuations resulting from VENC40 are lower than those from VENC120, implying the intra-aneurysmal flow data at VENC40 are more reliable than those at VENC120 in terms of the signal-to-noise ratio.

**FIGURE C1.**
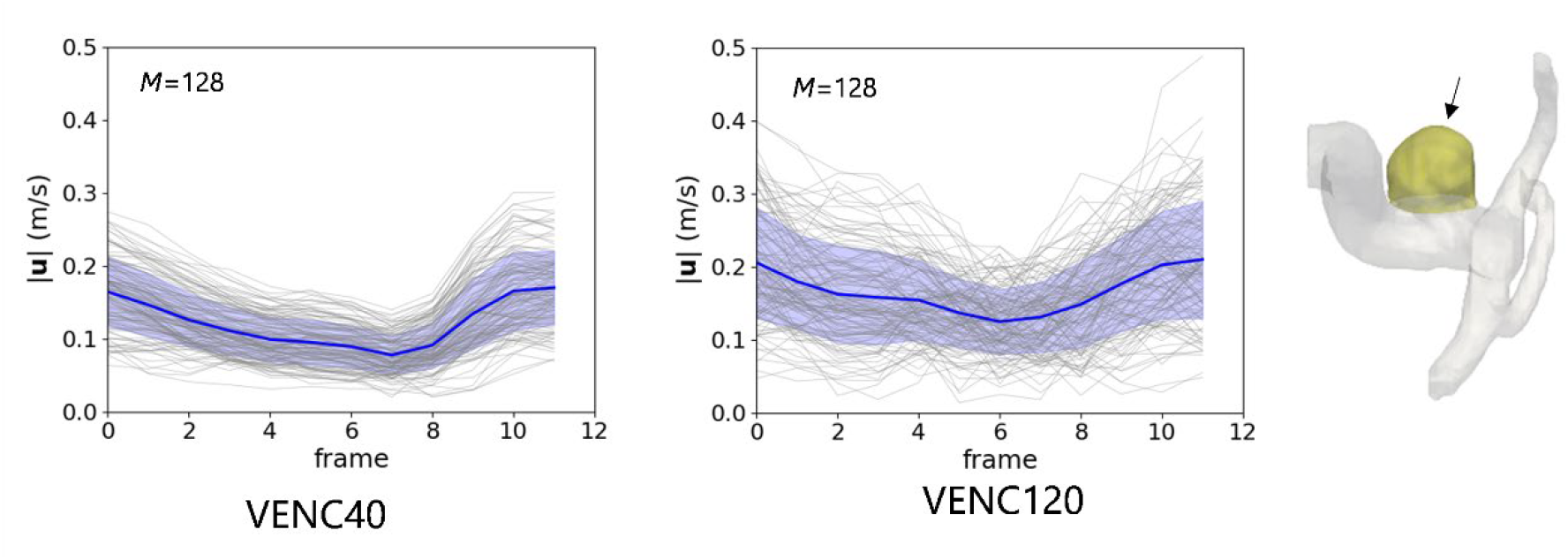
Temporal changes of the 4D flow MRI velocities in the aneurysm (colored by yellow in the right figure) at VENC40 and 120 (sample A).

